# Choroid plexus volume predicts expansion of chronic lesions and brain atrophy

**DOI:** 10.1101/2022.02.07.22270654

**Authors:** Samuel Klistorner, Michael H Barnett, Stuart L Graham, Chenyu Wang, Alexander Klistorner

**Affiliations:** Save Sight Institute, Sydney Medical School, University of Sydney, Sydney, New South Wales, Australia; Brain and Mind Centre, University of Sydney, Sydney, New South Wales, Australia; Sydney Neuroimaging Analysis Centre, Camperdown, New South Wales, Australia; Faculty of Medicine and Health Sciences, Macquarie University, Sydney, New South Wales, Australia

**Keywords:** chronic MS lesion, slow-burning inflammation, choroid plexus, lesion expansion

## Abstract

**Background and Objectives:** The expansion of long-standing multiple sclerosis (MS) lesions and an enlargement of choroid plexus are linked to chronic inflammation and microglial activation. In the current study, we investigated the association between plexus volume and subsequent lesion expansion in patients with relapsing remitting MS.

**Methods:** Pre- and post-gadolinium 3D-T1, 3D FLAIR and diffusion tensor images were acquired from 49 patients with relapsing-remitting MS. Choroid plexus volume and lesion activity were analysed between baseline and 48 months.

**Results:** Plexus volume remained stable during follow-up period. There was a strong correlation between baseline plexus volume and subsequent rate of chronic lesion expansion (r=0.77, p<0.001), which was stronger in close proximity to CSF. Furthermore, baseline plexus volume was also associated with change of Mean Diffusivity (MD) inside expanding area (r=0.55, p<0.001). There was, however, no correlation between baseline plexus volume and volume of new lesions. A cut-off of 98 × 10^−5^ plexus/TIV ratio predicted future lesion expansion with a sensitivity of 85% and specificity of 76%. Plexus volume larger than a cut-off was associated with >8-fold increased risk of chronic lesion expansion. Furthermore, baseline plexus volume significantly correlated with change of MD in lesional core during the study period (r=0.67, p<0.001) and with central brain atrophy (r=0.57, p<0.001).

**Conclusion:** Our data demonstrate that baseline plexus volume predicts subsequent expansion of chronic periventricular MS lesions and associated tissue damage.

## Introduction

Expansion of chronic MS lesions, caused by slow-burning inflammation at the lesion rim, has been implicated as one of the main forces driving MS progression.^1,2,3,4^ We recently demonstrated that lesion expansion is most noticeable in close proximity to ventricles, suggesting a link between slow burning inflammation and cerebrospinal fluid (CSF).^5^

It is well established that the choroid plexus, located within the ventricular system of the central nervous system (CNS), plays an important role in the implementation of the immune response within the CNS^6^. The plexus is essential for the production of cerebrospinal fluid (CSF)^7^, but also responsible for regulating entry of immune cells and specific molecules to the brain parenchyma, providing a gateway for antigen trafficking between the CSF and the vascular circulation.^8 9^

Several recent publications have demonstrated an association between plexus volume and the degree of inflammation in brain tissue, suggesting that plexus volume may serve as a surrogate marker for MS disease activity.^10 11 12^

Therefore, the aim of the current study is to investigate the association between plexus volume and subsequent lesion expansion in patients with relapsing remitting MS (RRMS).

## Methods

The study was approved by University of Sydney and Macquarie University Human Research Ethics Committees and followed the tenets of the Declaration of Helsinki. Written informed consent was obtained from all participants.

### Subjects

Forty nine consecutive patients with established RRMS, defined according to the revised McDonald 2010 criteria^13^, were enrolled in a longitudinal study. Patients underwent MRI scans and clinical assessment at 0 months, 12 months and 60 months. The main analysis was performed between 12 months (termed “baseline”) and 60 months (termed “follow-up”), while scans performed at 0 months (termed “pre-study scans”) were used to identify (and exclude) newly developed lesions at the start of the study (i.e. between 0 and 12 months).^3^

### MRI protocol and analysis

MRI was performed using a 3T GE Discovery MR750 scanner (GE Medical Systems, Milwaukee, WI). The following MRI sequences were acquired: Pre- and post-contrast (gadolinium) Sagittal 3D T1, FLAIR CUBE, diffusion weighted MRI. JIM 7 software (Xinapse Systems, Essex, UK) was used to segment individual lesions (using co-registered T2 FLAIR images). Expansion of chronic lesions was estimated using custom-build software, as previously described.^14^ Specific acquisition parameters and MRI image processing are presented in Supplementary material.

Choroid plexus was semi-automatically segmented on co-registered gadolinium-enhanced T1 images (Fig.1) using JIM 7 software (Xinapse Systems, Essex, UK) by a trained analyst (AK) at baseline and follow-up timepoints. The analyst ensured the correct contour was selected by the software and performed manual adjustments where needed. To reduce inter-subject variability CP volume was normalised by Total Intracranial Volume (TIV), as previously suggested^12^ (FreeSurfer software (http://surfer.nmr.mgh.harvard.edu/)). Therefore, CP/TIV ratio (x1000) will be referred in this paper as CP volume.

**Fig. 1.**
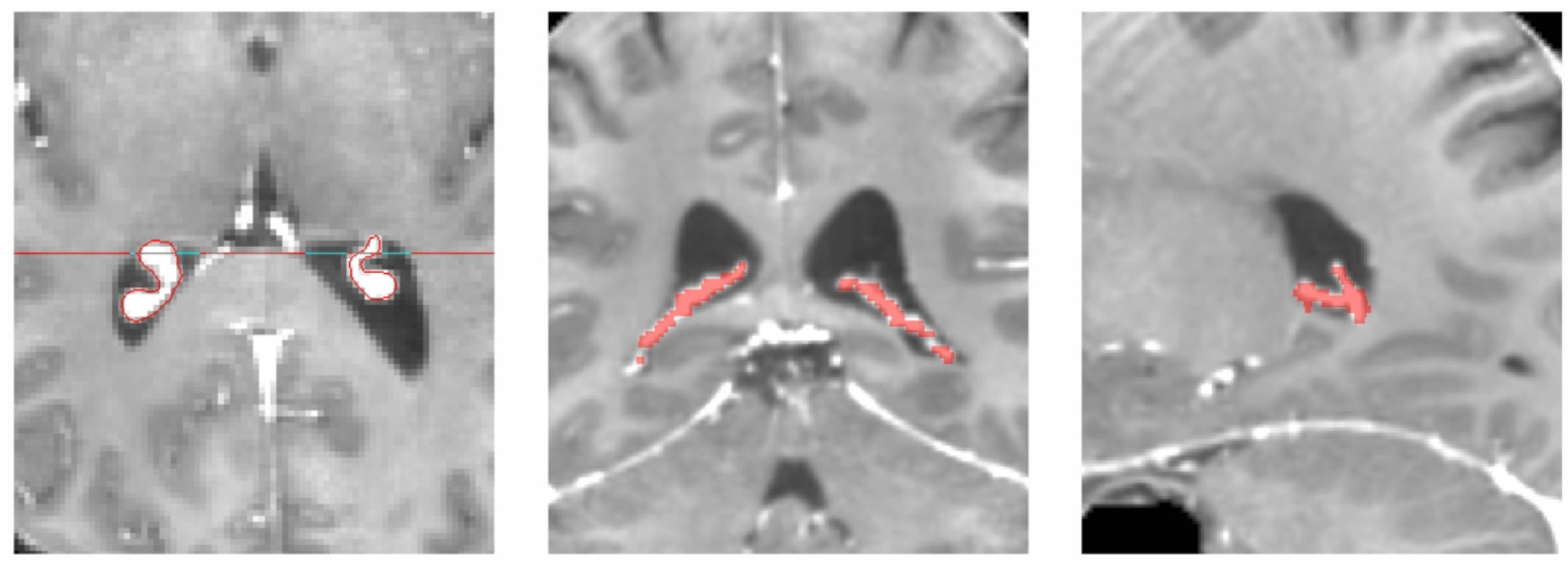
Example of plexus delineation using T1 GAD image. JIM9-assisted contouring was performed on axial image and verified using coronal and sagittal views.

To examine the effect of distance from CSF on the relationship between plexus volume and lesion expansion, periventricular brain tissue was separated into two 5 mm wide bands, one closer to CSF (1-5mm from ventricle) and one more remote (5-10mm) and the rate of lesion expansion and its relationship with plexus volume for each band measured independently. For band-based analysis, the number of voxels at baseline and follow-up within a single band were estimated for each patient in 3D space. The rate of expansion (percentage-wise) within the band was then calculated as a relative change in number of lesional voxels occurring during the follow-up.^3^

Volumetric change of lateral ventricles was used as a measure of central brain atrophy ^15 16^. It was calculated by multiplying baseline ventricle volume (as segmented by SIENAX) by the percentage ventricular volume change (as calculated by VIENA, a tool part of FSL).^17^

The degree of lesional tissue damage was assessed by Mean Diffusivity (MD). ^18 19^ Progressive tissue destruction in chronic lesions was measured as an increase of MD between baseline and follow-up timepoints^19 20^ using the baseline lesion mask, which was adjusted to correct for brain atrophy-related displacement of lesions at follow-up, as described previously. ^3^

### Statistics

Statistical analysis was performed using SPSS 22.0 (SPSS, Chicago, IL, USA). Pearson correlation coefficient was used to measure statistical dependence between two numerical variables. For partial correlation, data was adjusted for age, gender, disease duration, baseline lesion volume and ventricular size. P < 0.05 was considered statistically significant. Shapiro–Wilk test was used to test for normal distribution. Comparisons between groups were made using Student *t*-test. Longitudinal changes were assessed using paired two-sample *t*-test. Fisher’s exact test was used for categorical data. To investigate how well plexus volume predicts tissue loss inside chronic lesions and brain atrophy after 4 years of follow-up, we performed a forward stepwise linear regression analysis that included age, gender, disease duration, baseline ventricular volume, baseline lesion volume and volume of new (confluent and free standing) lesions. Receiver operating characteristic (ROC) curve analysis was performed to determine sensitivity and specificity of baseline plexus volume in predicting future lesion expansion, while hazard ratio was calculated to estimate relative risk of chronic lesion expansion in patients who demonstrated plexus enlargement.

## Results

Forty-nine consecutive MS patients enrolled in longitudinal study of axonal loss in MS who reached 5 years follow-up were included in the study. 10 patients remained on low-efficacy treatment (injectables, such as interferon and glatiramer acetate, teriflunomide and dimethyl-fumarate)^21^ and 21 patients were receiving high-efficacy drugs (fingolimod, natalizumab, and alemtuzumab)^21^ during the study period. Two patients were treatment-free, while 16 patients changed treatment category between baseline and follow-up visits. Demographic and lesion data in presented in Table 1.

**Table 1.**
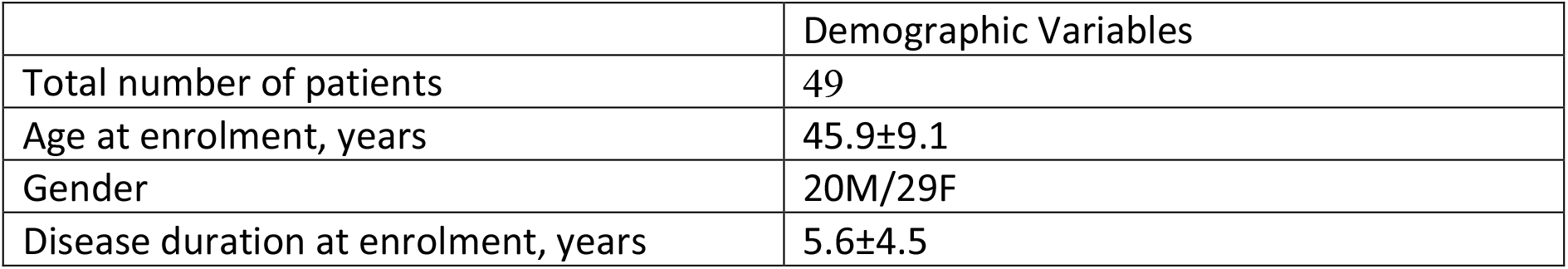
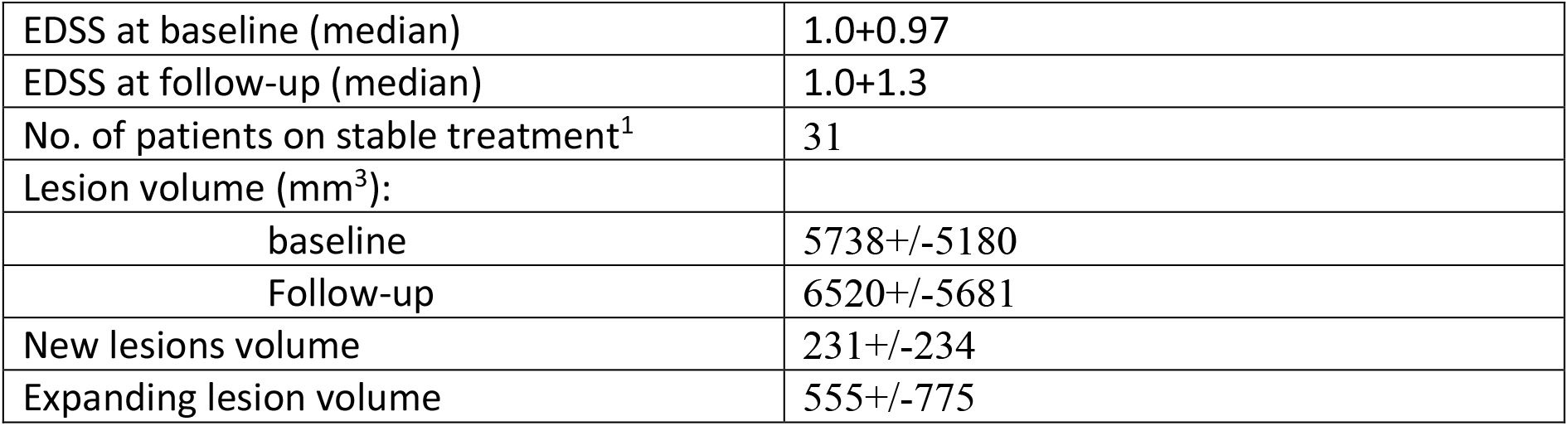
Demographic data

Example of plexus delineation is shown in Fig. 1.

No correlation was found between baseline plexus volume and baseline lesion volume (r=0.25, p=0.08), ventricle volume (r=0.24, p=0.1), gender (r=0.16, p=0.27 Spearman) or disease duration (r=0.08, p=0.6). There was also no significant difference in baseline plexus volume between the two treatment groups (87.4 and 101 respectively, p=0.3). Plexus volume, however, was significantly associated with baseline EDSS (r=0.44, p=0.01) and patient age (r=0.32, p=0.02).

There was no significant change of plexus volume during follow-up (p=0.45, paired t-test).

### Baseline plexus volume predicts subsequent expansion of chronic lesions

We observed a strong association between baseline plexus volume and subsequent rate of lesion expansion (r=0.77, p<0.001). This correlation remained materially unchanged after adjusting for age, gender, disease duration, ventricular volume and baseline lesion volume (partial correlation r=0.74, p<0.001) (Fig.2). Furthermore, plexus volume correlated with change of MD inside expanding area (r=0.55, p<0.001).

**Fig. 2.**
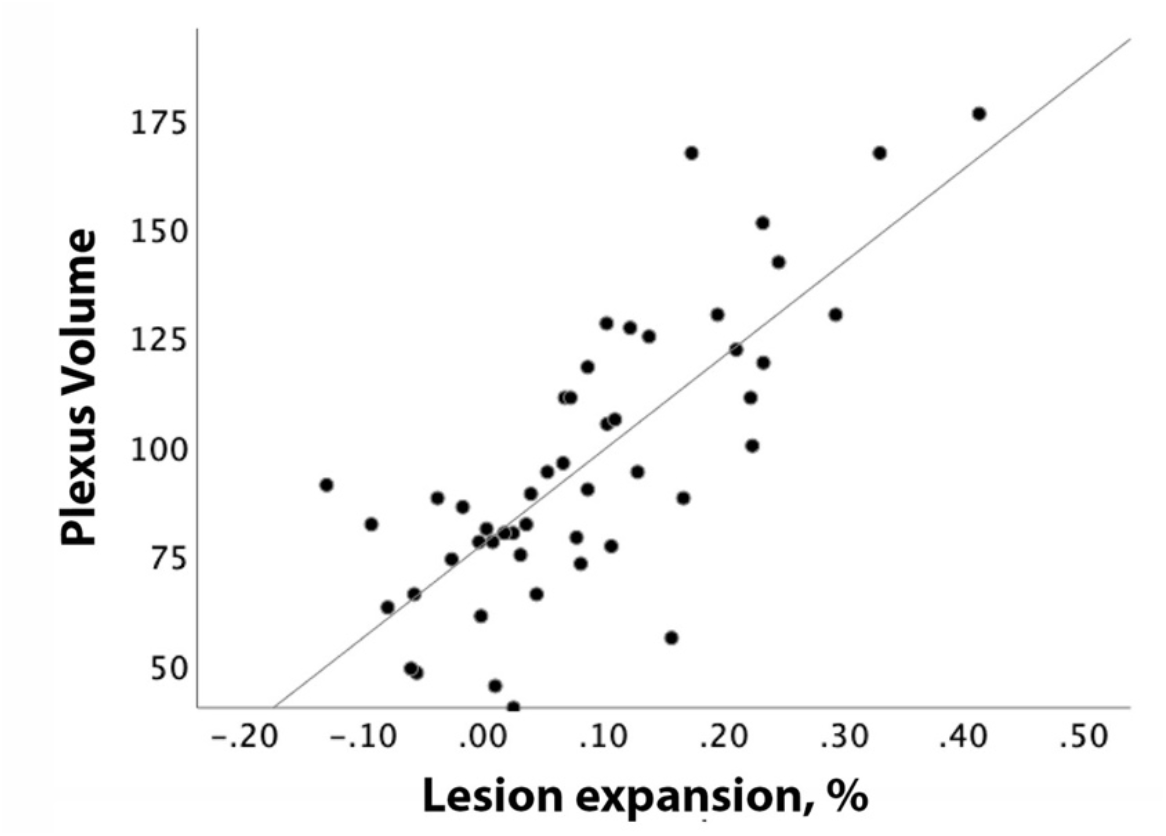
Correlation between baseline plexus volume and patient-based percentage of chronic lesion expansion.

There was, however, no significant correlation between baseline plexus volume and volume of new (combined confluent and free-standing) lesions (r=0.08, p=0.5, partial correlation r=0.011, p=0.4).

Alteration of CP/TIV ratio during follow-up did not correlate with volume of new lesions or the rate of chronic lesion expansion (r=0.01, p=0.9 and r=0.14, p=0.4).

Patient-based receiver operating characteristic (ROC) curve analysis of baseline plexus volume (area under the curve = 0.89) yielded a cut-off of 98 × 10^−5^ plexus/TIV ratio, which predicted future lesion expansion with a sensitivity of 85% and specificity of 76% (Fig.3).

**Fig. 3.**
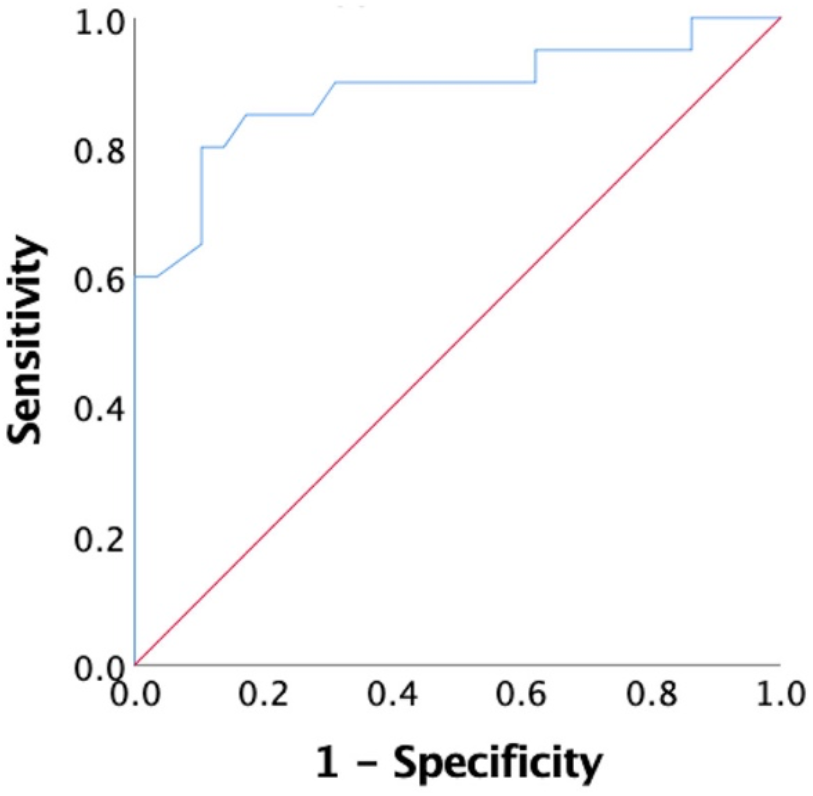
ROC curve showing sensitivity and specificity of baseline plexus volume in detecting expansion of chronic lesions

Furthermore, based on degree of chronic lesion volume increase, patients were separated into two groups: patients who demonstrated significant (i.e. >10%) lesion expansion and patients with stable chronic lesion volume (lesion expansion <10%)^14^. Plexus volume larger than a cut-off calculated using ROC analysis (i.e. 98 × 10^−5^ plexus/TIV ratio) was associated with >8-fold increased risk of chronic lesion expansion [hazard ratio for lesion expansion comparing enlarged vs non-enlarged plexus = 8.4; 95% confidence interval (CI): 2.8 to 25.0).

### Plexus volume correlates with tissue damage inside MS lesions and loss of extra-lesional brain tissue

We have previously ascribed the association between expansion of chronic lesions and lesional or brain tissue loss to axonal and neuronal death following transection of axons at the expanding rim caused by slow-burning inflammation^3 20^. Therefore, to investigate whether individual differences in plexus volume translate to neuro-axonal loss, we examined the potential association of plexus volume with markers of lesional tissue damage (as assessed by an increase of Mean Diffusivity, MD)^19^ and brain atrophy.

Plexus volume at baseline demonstrated a significant correlation with baseline MD in lesional core (r=0.58, p<0.001, partial correlation adjusted for age, gender, duration, ventricular volume and baseline lesion volume, r=0.42, p=0.004), which increased at follow-up (r=0.66, p<0.001, partial correlation r=0.60, p<0.001).

Remarkably, baseline plexus volume also significantly correlated with change of MD in lesional core during the study period (r=0.67, p<0.001, partial correlation, 0.66, p<0.001 respectively). Furthermore, baseline plexus volume also showed a similar association with ventricular enlargement: (r=0.57, p<0.001, partial correlation: r=0.51, p<0.001). There was, however, no association observed between plexus volume and EDSS change during follow-up.

Considering age-related progression of ventricular expansion in healthy subjects (0.8% per year)^22 23^, patients were separated into two groups: progressors (i.e. patients with >3.2% ventricular expansion in 4 years) and non-progressors (<3.2% ventricular expansion in 4 years). There was significantly larger baseline plexus volume in the progressors group compared to non-progressors (101.1+/-32.4 vs 77.6+/- 24.2 respectively, p=0.01) (Fig.4).

**Fig. 4.**
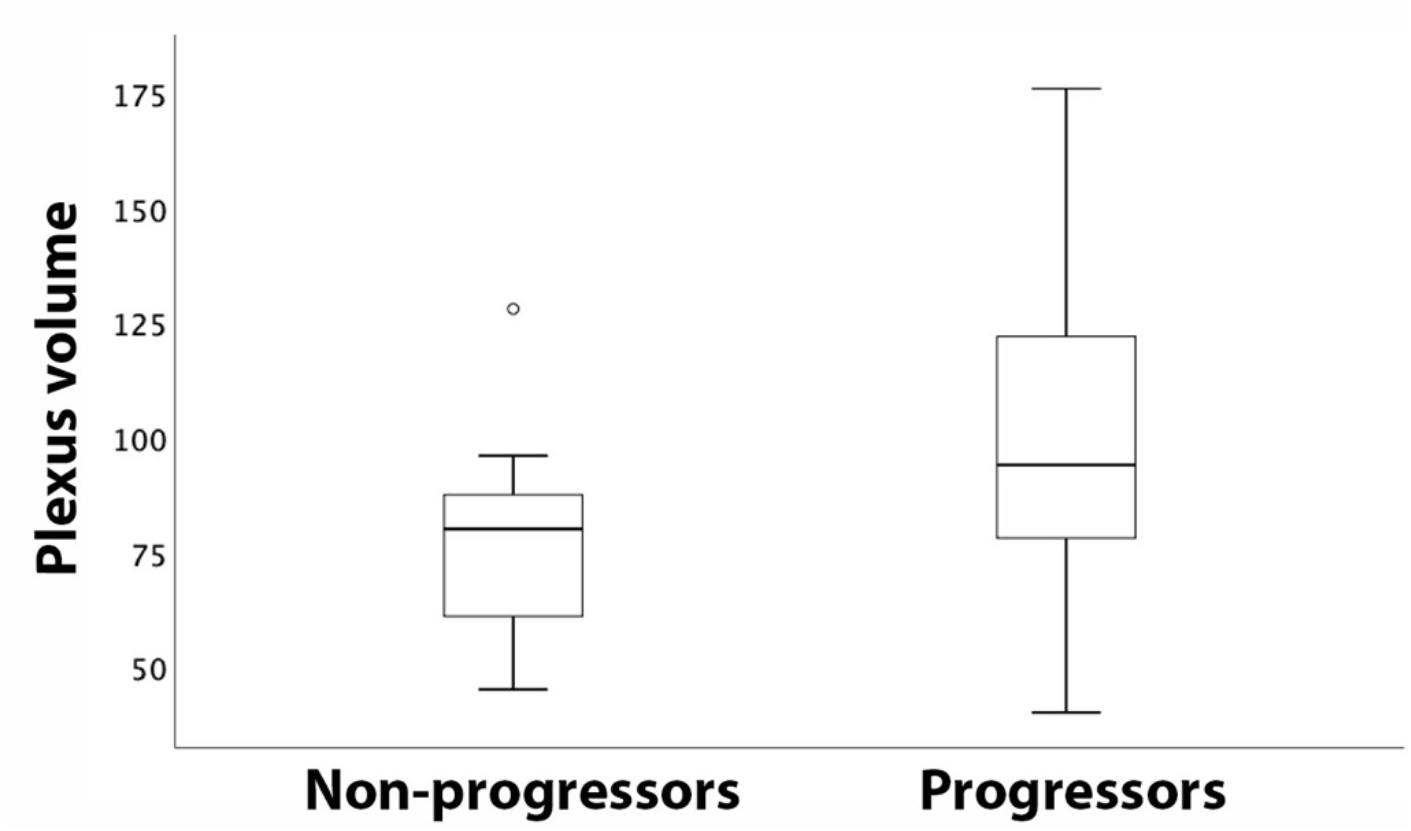
Box-plot of plexus volume in patients with stable and expanding ventricles

In accordance with our previous publications^19 14^, we found a significant correlation between volume of lesion expansion and baseline MD in the lesional core (r=0.47, p<0.001, partial correlation adjusted for age, gender, duration r=0.40, p=0.005), which also increased at follow-up (r=0.61, p<0.001, partial correlation adjusted for age, gender, duration, r=0.57, p<0.001). Moreover, expansion of chronic lesions correlated with change of MD during the study period (r=0.63, p<0.001, partial correlation adjusted for age, gender, duration, r=0.62, p<0.001 respectively). Similarly strong relationships were observed between lesion expansion and brain atrophy (r=0.71, p<0.001, partial: r=0.67, p<0.001).

### Linear regression model

To investigate further the impact of plexus volume on predicting future tissue loss inside (MD change) and outside (brain atrophy) chronic lesions, a linear regression model was performed.

Change of MD inside chronic lesions (model R^2^=0.51, p<0.001) was driven almost entirely by baseline plexus volume (Standardized coefficient Beta: 0.62, p<0.001 and 0.25, p=0.02 for plexus volume and volume of new lesions respectively). Other variables did not reach significance (Fig 5a). Removal of baseline plexus volume had a dramatic effect on the model predictive power (R^2^=0.13, p=0.01, F change <0.001)

**Fig. 5.**
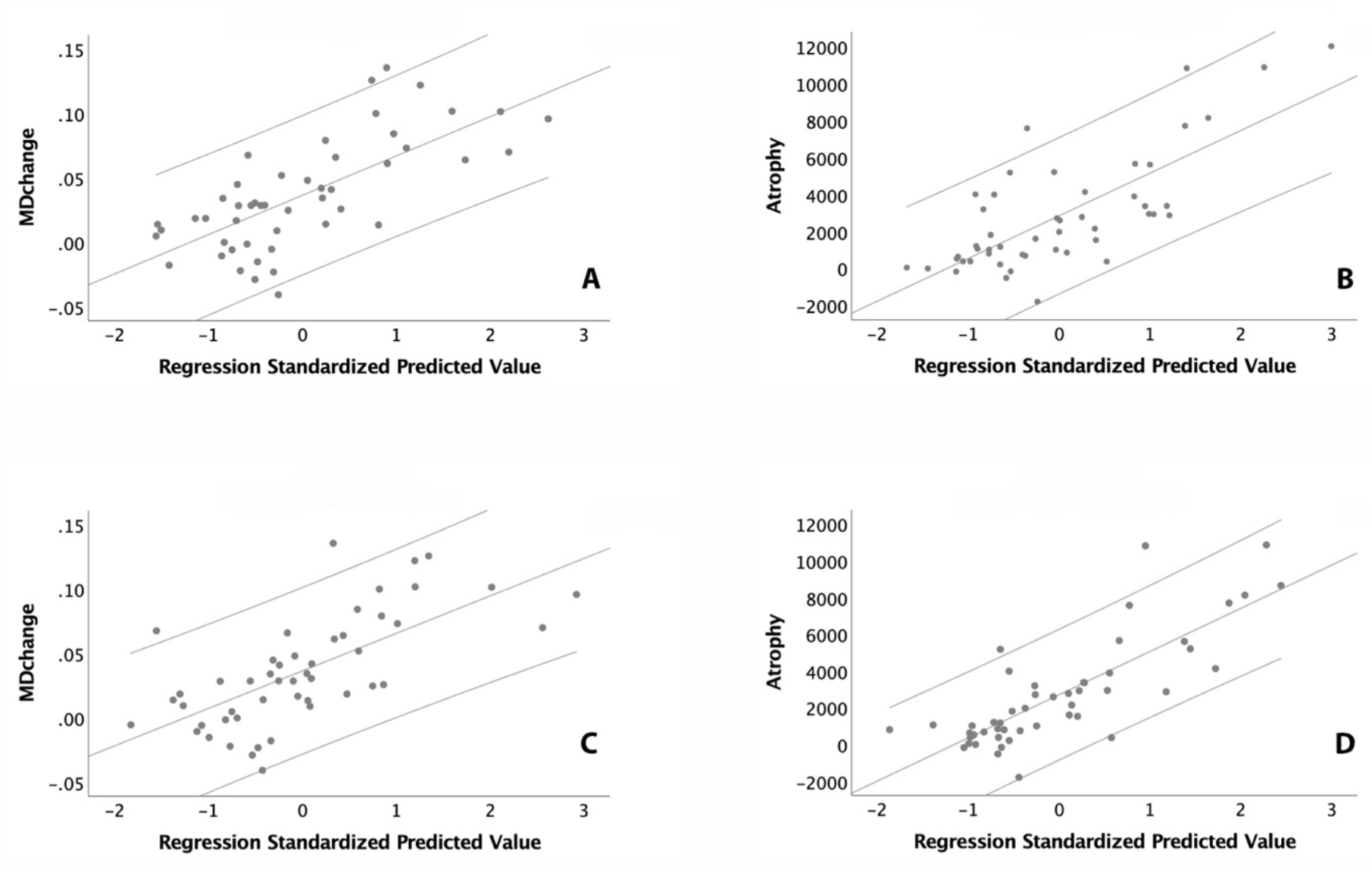
a, b. Linear regression analysis of baseline plexus volume vs lesional MD (a) and brain atrophy (b) c, d. Linear regression analysis of expansion of chronic lesions vs lesional MD (c) and brain atrophy (b)

For the atrophy model (R^2^=0.56, p<0.001), baseline plexus volume was also the main contributor to the tissue loss outside of lesions (Standardized coefficient Beta: 0.44, 0.35 and 0.31 for plexus volume, volume of new lesions and baseline ventricular volume respectively, p<0.001, = 0.001 and 0.003 respectively) (Fig. 5b). Again, exclusion of baseline plexus volume significantly reduced the predictive power of the model (R^2^=0.37, p<0.001, F change <0.001).

The addition of lesion expansion as a dependent variable to progressive lesional MD change or brain atrophy models revealed a high degree of collinearity between the lesion expansion rate and baseline plexus volume (variance proportions 0.98 and 0.56 respectively). As a result, substitution of plexus volume with rate of chronic lesion expansion did not materially change the model’s predictive power. Thus, using lesion expansion during the follow-up period rather than baseline plexus volume for modelling MD change in the lesional core demonstrated predictive power of R^2^=0.46, p<0.001 and was driven by the volume of lesion expansion and new lesions (Standardized coefficient Beta: 0.58, p<0.001 and 0.26, p=0.02 for lesion expansion and new lesions respectively) (Fig.5 c). Modelling brain atrophy using similar substitution revealed R^2^=0.63 (p<0.001), with lesion expansion, volume of new lesions and ventricular volume showing significant contribution (Standardized coefficient Beta: 0.67, 0.26 and 0.25, p<0.001 0.007 and 0.015 respectively) (Fig. 5d).

### Correlation between plexus volume and lesion expansion is related to the distance from CSF

We measured the rate of lesion expansion and its relationship with plexus volume in the two bands independently. While plexus volume explained 50% of the variability in the rate of lesion expansion in the band closer to ventricles, this relationship was significantly weaker (p=0.01) for the more distal band (r=0.71, p<0.001 vs r=37, p=0.01 in close and remote rings respectively) (Fig.6).

**Fig. 6.**
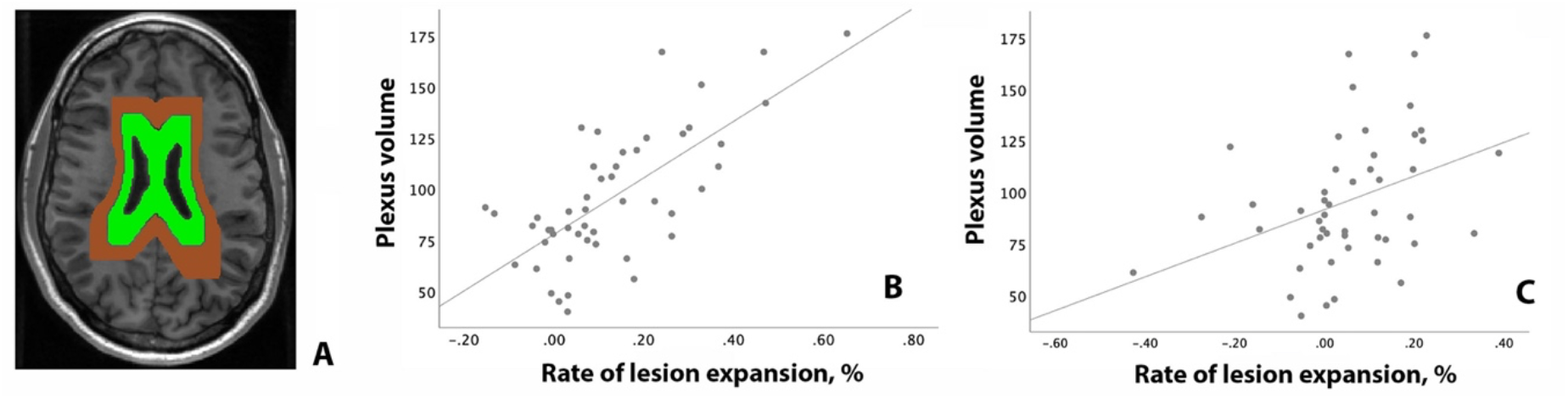
Analysis of association between plexus volume and chronic lesion expansion at different distance from ventricles. a. Example of two-bands mask b. Correlation between plexus volume and lesion expansion in the band adjacent to ventricles (green) c. Correlation between plexus volume and lesion expansion in the more distal band (brown)

## Discussion

### Plexus volume correlates with subsequent expansion of chronic lesion

Slow-burning inflammatory demyelination at the edge of chronic MS lesions (and lesion expansion as its imaging equivalent) has been implicated as a major factor contributing to disease evolution, including progressive neurodegeneration, brain atrophy and disability worsening.^1 2 3^ Here we explored the relationship between the rate of expansion in chronic MS lesions and volume of choroid plexus in sizable cohort of RRMS patients. We demonstrated a strong association between the size of the choroid plexus at baseline and subsequent expansion of chronic lesions. This relationship remained unchanged after adjustment was made for age, gender, disease duration, ventricular volume and baseline lesion volume. A close link between baseline plexus volume and lesion expansion was also supported by the observation that the association between the two is stronger in close proximity to the ventricles, where the plexus resides. Furthermore, the predictive value of plexus volume in identifying future lesion expansion was corroborated by the ROC analysis, which revealed that a cut-off of 98 × 10^−5^ plexus/TIV ratio predicted which patients would exhibit future lesion expansion with a sensitivity of 85% and specificity of 76%. Patients with baseline plexus volume above this threshold had an 8-fold increased risk of chronic lesion expansion. In addition, degree of tissue loss within expanding part of chronic lesions (as measured by increase of MD) was also positively linked to the size of the plexus at baseline.

Surprisingly, we found that plexus/TIV ratio remains stable during 4 years of follow up, suggesting that plexus enlargement occurs early in the course of MS, which is supported by recent study of Ricigliano et al.^12^ Stability of plexus volume and its weak association with baseline lesion volume and with the volume and severity of new lesions implies that, at least in our cohort, increased plexus size relates to a pathogenesis that is discrete from acute inflammatory demyelination, supporting, therefore, the notion that different mechanisms are likely to be responsible for acute and smouldering inflammation.^1 2 14 24 25^

The essential role of the choroid plexus in maintaining the blood–CSF barrier and modulation of inflammatory cells trafficking into the central nervous system has recently attracted the attention of MS researchers.^26 27^ It has become clear that the plexus not only actively participates in acute inflammatory processes including antigen presentation and recruitment of peripheral inflammatory cells^28^, but remains chronically inflamed, even in long-standing MS^8^.

Several recent studies have examined the size of the choroid plexus in MS patients and its potential relationship with disease progression. Ricigliano et al^12^ demonstrated a larger volume, and increased inflammation, of the choroid plexus compared with normal controls, particularly in the relapsing-remitting stage of the disease. The authors also reported an association of larger plexus size with measures of disease activity, including relapse rate or the number of enhancing lesions, and parenchymal neuroinflammation. A close relationship between enlargement of choroid plexus and neuroinflammation in both MS and experimental models of demyelination (cuprizone) and experimental autoimmune encephalomyelitis (EAE) has also been demonstrated in another study^10^ in which choroid plexus volume outperformed conventional MRI biomarkers in predicting EDSS progression.

Various hypotheses have been proposed to explain plexus enlargement in MS. Inflammation through activation and infiltration of immune cells^26^, endothelial immune proliferation, increase of permeability of the Blood-CSF Barrier^10^, CSF hypersecretion^29^, oxidative stress^30^ and edema^31^ have all been suggested as potential contributing factors. Furthermore, it was recently demonstrated that plexus volume is related to the increased activity of key innate immune cells in the CNS, namely astrocytes and microglia^10^.

Interestingly, microglial activation also plays a central role in sustaining slow-burning inflammation at the rim of chronic lesions,^32 33 34 35 36^ which may suggest a shared pathomechanism between plexus inflammation and lesion expansion. Thus, for instance, it is feasible that soluble cytotoxic factors and/or inflammatory mediators produced by long-standing intrathecal inflammation (including choroid plexus, but also meningeal infiltrates and interstitial fluid^9^) could diffuse from CSF into the periventricular white matter, forming /sustaining a permanent inflammatory environment in and around the ventricles, thereby driving slow-burning inflammation at the rim of chronic periventricular lesions.^37 38 39^

This concept is supported by the recently described periventricular gradient of chronic lesion expansion, which demonstrated a decrease in a rate of chronic lesion enlargement as distance from the CSF (and plexus) increases^5^; and by the observation reported in this study that the association between baseline plexus volume and lesion expansion is stronger in close proximity to the CSF. Notably, the periventricular gradient of lesion expansion is strikingly similar to the pattern of microstructural damage in the white matter described near the ventricles and in the most external cortical layers (i.e. adjacent to CSF-filled ventricular and subarachnoid spaces), which is believed to be mediated by innate immune cell activation.^40 41 38 39^ This common mechanism of inflammation, found in both CSF and meninges^42 9^, has recently prompted a reappraisal of the pathogenesis of MS, particularly of its chronic progressive stage.^43^

While the possibility that perilesional inflammation causes diffusion of oxidative factors from the brain parenchyma into the CSF compartment, reaching the CP epithelium and leading to CP inflammation and expansion can not be completely eliminated^27^, our longitudinal data suggest that this “reverse causation” is unlikely.

### The association between CP volume and brain tissue loss inside lesions and in NAWM is likely to be mediated by rim inflammation

We previously reported a close association of chronic lesion expansion with progressive increase of MD inside established lesions and with brain atrophy; this finding was replicated in the current study. Progressive tissue damage inside expanding lesions has also been reported by others groups.^2 44^ The mechanism of MD change inside chronic lesions was attributed to widening of the extracellular space following Wallerian and retrograde degeneration of the intra-lesional part of axons transected at the lesion rim (as a result of slow-burning inflammation).^20 14^ Similarly, degeneration of the extra-lesional component of the same axons leads to the elimination of their associated axolemmae and myelin sheaths from the white matter, neuronal death and ultimately to brain atrophy.

The current study demonstrated that baseline plexus volume also significantly correlates with progressive change of isotropic water diffusion (MD) inside chronic lesions and with brain atrophy (as measured by ventricular enlargement) over 4 years of follow-up. In addition, group analysis demonstrated a significantly higher volume of choroid plexus in patients with progressive brain atrophy compared to “non-progressors”.

However, considering the strong correlations between baseline plexus volume and subsequent chronic lesion expansion on one hand, and between the latter and measures of tissue loss on the other, we hypothesise that the observed association between plexus metrics and progressive change in lesional MD or brain atrophy is likely to be an indirect consequence of the relationship between the plexus and lesion expansion.

Furthermore, the high collinearity and virtually interchangeable character of baseline plexus volume and subsequent expansion of chronic lesions in models predicting progressive brain damage (as shown by Linear Regression analysis) also lends strong support to the notion that, while baseline plexus volume explains about 50% of the variability in tissue damage both within and outside of chronic lesions in following four years, the mechanism of this association is likely to be operating via inflammatory activity at the rim of chronic lesions and accompanying lesion expansion.

The limitations of our study are a relatively small sample size, heterogeneity of disease modifying therapies and the absence of normal controls. However, the incorporation of normal controls, who lack demyelinating lesions or atrophic changes, would not influence our findings. Furthermore, a comparison with normal subjects has been performed in several earlier studies.

We were not able to demonstrate a correlation between plexus volume and EDSS change, reflecting a lack of progression of mean EDSS over the course of the study. This may reflect a lack of progression of mean EDSS over the course of the study or insensitivity of the EDSS to periventricular lesional damage.

In summary, our study demonstrated strong correlations of choroid plexus size with: 1) subsequent expansion of chronic periventricular MS lesions; 2) associated axonal loss within established lesions; and 3) brain atrophy. While the molecular mechanisms underpinning these relations require further investigation, the longitudinal nature of the study, the strength of the reported associations and higher strength of correlation between plexus volume and lesion expansion in close proximity to CSF/plexus, together with observed temporal sequence of events argue in favour of a causal relationship between the volume of choroid plexus and subsequent lesion expansion.^45^

## Data Availability

All data produced in the present study are available upon reasonable request to the authors

## Notes

### Competing Interest Statement

The authors have declared no competing interest.

### Funding Statement

This study did not receive any funding

